# Childhood Ocular Inflammation Sensations and Symptoms Reporting Questionnaire (ChOIR-Q): Development and assessment of a paediatric self-report tool

**DOI:** 10.1101/2024.04.06.24305169

**Authors:** Ameenat L Solebo, Salomey Kellett, Jugnoo Rahi, Andrew D Dick, Jane Ashworth, Gisella Cooper, Eibhlin McLoone, Kirithika Muthusamy, Rachel Pilling, Harry Petrushkin, Valerija Tadic

**Affiliations:** Population, Policy and Practice Department, UCL GOS Institute of Child Health, London, UK; Ophthalmology Department, Great Ormond Street Hospital, London, UK; Great Ormond Street Hospital Biomedical Research Centre, London, UK; Ulverscroft Vision Research Group, London, UK; Rheumatology Department, Great Ormond Street Hospital, London, UK; UCL Institute of Ophthalmology, London, UK; Moorfields Biomedical Research Centre, London, UK; School of Cellular and Molecular Medicine, Bristol University, Bristol UK; Paediatric Ophthalmology, Manchester Royal Eye Hospital, Manchester, UK; Division of Evolution, Infection and Genomics, University of Manchester, Manchester, UK; Ophthalmology Department, Sheffield Children’s Hospital, Sheffield, UK; Ophthalmology Department, Belfast Children’s Hospital, Belfast, UK; External disease Department, Moorfields Eye Hospital, London, UK; School of Optometry and Vision Science, University of Bradford, Bradford, UK; Paediatric Ophthalmology Department, Bradford Royal Infirmary, Bradford, UK; Uveitis Department, Moorfields Eye Hospital, London, UK; School of Human Sciences, Greenwich University, London, UK

**Keywords:** Patient Reported Outcome Measures (PROMs), Health Measurement Scales, Ocular

## Abstract

We aimed to develop and assess age-appropriate child and young person, self and proxy report tools to capture and characterise eye symptoms in childhood ocular inflammatory disease.

Children and young people aged under 18 years diagnosed with inflammatory eye disease (uveitis), and their families, were recruited to a multiphase study, involving: text and pictogram items generation through focus groups and interviews (Phase 1), pre-testing face validity analysis including and discussion amongst a multidisciplinary professional panel (Phase 2), and pre-piloting (Phase 3) and piloting (Phase 4) of the instrument amongst a representative sample of the target population.

A total of 170 participants, comprising 113 children/young people and 57 parents/carers, were recruited. Phase 1 resulted in the generation of 60 items. Following phases 2 to 3, these items were developed into self-completion, and assisted self-completion tools for children aged 9 years and older, and those aged under 9 years respectively, and a proxy score, for completion by parents and carers. Correlations scores between individual item and whole domain scoring were above 0.58 for the self-completion tools and above 0.39 for the proxy completion tool. Initial Cronbach’s alpha for the tool overall was good at 0.84, with within-domain alphas of 0.81 – 0.87.

In conclusion, these instruments demonstrate the feasibility of capturing ocular sensations in children and young people, with a patient centred development approach resulting in tools with high rates of completion, and acceptable internal instrument consistency.

## 1. Introduction

Childhood ocular inflammatory disorders, an otherwise heterogenous group of conditions, are collectively characterised by frequent co-occurrence of multi-system inflammatory disorders such as juvenile idiopathic arthritis, sarcoidosis, Behcet’s and inflammatory bowel disease.^1–3^ These eye diseases and associated disorders confer negative impact on vison and quality of life, with accumulation of years lived with subsequent morbidity.^1^ As with other headache and facial pain, the challenge in articulating ocular sensations and symptoms can be an obstacle to accessing the appropriate health professionals needed to secure the ocular diagnosis, with timely diagnosis being a key predictor of better visual outcomes. Timely diagnosis ocular inflammation also enables prompt diagnosis and treatment of underling systemic disorders.

An exemplar childhood onset ocular inflammatory disorder is uveitis.^4^ Uveitis is currently estimated to affect one in 1,000 children,^5,6^ It often co-exists with other inflammatory eye disorders such as scleritis, keratitis or orbital inflammation.^7–11^ Disease and treatment can involve or impact all areas of the eye.^1^ In the early stages the most common form of disease (a chronic anterior form of uveitis) is often described as asymptomatic, in striking contrast to the ‘explosively’ painful and red acute-onset phenotype of similar adult disease.^12,13^ This description may be true, or may, more likely, reflect challenges in capturing a child’s experience of ocular sensations and symptoms with the more insidious presentation seen in early life. Uveitis onset is bimodal, most commonly presenting at age 2–5 years, and another smaller peak in adolescence.^12,14^ Younger children presenting with incident disease or with recurrences may struggle to articulate ocular sensations, lacking the necessary cognitive maturity or vocabulary,^15^ with reports of initially asymptomatic cases of childhood uveitis having symptomatic presentation in later childhood.^16^ Older children may also struggle to describe ocular sensations. There are no validated child-centred tools to capture the experience of eye sensations or symptoms in young children with ocular inflammatory disorders. There is also an absence of available adult measures which might be adapted, with appropriate consideration of differences, for use by young people.^17^

A reliable, repeatable patient-reported measure (e.g., with high face and content validity)^18^ of the negative ocular sensations and symptoms would be a valuable disease management tool, capturing information that may indicate underlying aetiology when used as a surveillance or triage tool, or one which complements clinical assessment when used in disease monitoring. We report the development and early assessment of age-appropriate child, young person and family, self and proxy reporting tools which aim to capture and characterise ocular symptoms in childhood ocular inflammatory disease.

## 2. Methods

### 2.1. Study design

This was a multiphase, multicentre, prospective study. The COSMIN (COnsensus-based Standards for the selection of health Measurement INstruments) Study Design checklist for Patient-reported outcome measurement instruments,^19^ the PROMIS (Patient-Reported Outcomes Measurement Information System) initiative recommendations,^20–22^ and the consolidated criteria for reporting qualitative research (COREQ) checklist^23^ were used to develop study methods. The phases comprised: item generation for the instrument (Phase 1), pre-testing face validity analysis (Phase 2), pre-piloting and piloting of the instrument (Phases 3 and 4). Development and assessment of the tool was supported by a patient expert group (the Childhood Uveitis Study Steering Group) and assessment of the tool was further supported by a professional group (the Childhood Ocular Inflammation Sensations and Symptoms Reporting Questionnaire, or ChOIR-Q Professional Group, JA, GC, LS, EMc, EMo, KM, RP, HP).

### 2.2 Patient identification and recruitment

Children / young people aged under 18 years diagnosed with uveitis were eligible. Those with a neurological or developmental impairment which was considered (by their parents / carers) to preclude involvement were excluded from recruitment as interviewees to the qualitative phases of the study. Participants were recruited through advertisements placed in UK hospitals, or on attendance to the Uveitis service within Great Ormond Street or Moorfields Eye Hospitals. Those aged 16 to 18 years gave informed consent for participation with parental consent given for those under 16.

### 2.3 Study Phases

#### Phase 1: item generation

Six focus groups (3-4 children with one parent/carer each, mixed age and mixed gender) and six one-to-one interviews were conducted by an experienced moderator / interviewer (ALS), with an experienced moderator (VT) present for the first focus group. Purposive sampling was undertaken to ensure representation across age groups, sexes and ethnic backgrounds. Age groups were categorised in line with consensus based guidance from the patient expert group as pre/early verbal children, 2 to under 5 years old; young children, 5 to 8 years; ‘peri-pubertal’ children, 9 to 12 years; and adolescents / young people aged 13 years and over).

Group and interview discussions were undertaken using a semi-structured interview topic guide (supplement), developed using existing validated general paediatric pain scores.^21,24–28^ This included the pre-identified domains of the characteristics, severity, alleviators and exacerbators, location, temporality, and external presentation of ocular sensations and symptoms. Photo-elicitation (incorporation of images into interviews) involved banks of images representing negative ocular sensory imagery as identified by the investigators. Whilst topic guide structure was unchanged between focus groups, comments from previous groups were used to inform subsequent group moderation, with the creation of new prompts and images after each focus group. Each participant received a £20 gift card for their contribution.

Focus groups and interviews were audio recorded and transcribed. Content analysis was undertaken following each group or interview, to identify potential items and to allocate them within the existing core domains (characteristics, severity, alleviators and exacerbators, location, temporality, external presentation, and proxy items) where possible. Draft items that did not map to any of these domains, were coded as ‘Other’ and again informed the moderation of the subsequent focus group or interviews. Content analysis on responses around imagery informed the creation and iterative refinement of novel study specific image pool items. A summary report of participant opinions on the utility or likeliness of using the proposed instrument was compiled.

#### Phase 2: Pre-testing and establishing face validity

The items generated by Phase 1 were tested with a different and wider participant group through an online survey (built within REDCap) to determine comprehensibility and relevance (face validity). The survey invitation, accompanying explanatory document, and QR link for the survey, were distributed within collaborating paediatric uveitis centres. Respondents were asked to report similarity between terms and suggest item rephrasing and select preferred modes of administration. Descriptive analyses of responses from all participants were undertaken, with subgroup analyses of responses categorised by age of child: any item not deemed applicable or intelligible, or considered to be replicating another item by at least two carers or children within any of the age bands (aged 13 years and over, 5 - 12 years, under 5 years old) was flagged for removal from the potential item pool.

To further establish item relevance and face validity, items and survey responses were shared with professional and patient stakeholders at a ‘Council of expertise’ (ChOIR-Q Professional Group) with members representing multi-disciplinary professionals, including ophthalmologists managing non-uveitis childhood ocular inflammatory disorders (orbital inflammation, keratitis, scleritis and optic neuritis) from high and low volume clinical centres. Items flagged for removal from the item pool by the patient survey (0% voted as relevant for childhood ocular disease) were discussed amongst the council, and any items which the majority (>50% of the council) considered to be indicative of disease other than uveitis were noted. The council were asked to comment on ‘best fit’ terms selected for use where the patient survey had indicated high similarity of terms (term similarity noted by more than 2 respondents), and to comment on the modification of retained items (modified to adhere to PROMIS development team requirements, e.g., consistency of tenses, standard response options).^21^

#### Phase 3: Tool pre-piloting

The three resultant draft instruments (a child self-completion tool, child self-completion with parental help tool, and a parent or carer proxy completed tool) were administered in person or over telephone to a consecutive convenience sample of patients attending the Uveitis Service at Great Ormond Street and Moorfields Eye Hospitals, as a baseline population for the framework. At least 3 participants were recruited in each age-group (total n=12 patients).

Children, young people (for self-report) and carers (for proxy report for children aged under 5 years) completed one tool independent of the researcher during the 24 hours before their routine follow up clinic appointment. Following a structured interview, a summary statement for each item compiling the young person’s, child’s or the parent and carer’s comments was created. The tool development team then reviewed comments to reach final agreement on item order, instructions, and response options.

#### Phase 4: Piloting

The new instruments were tested on a larger representative sample to judge instrument feasibility and acceptability. Participants were recruited consecutively on attendance to the Uveitis Service at Great Ormond Street and Moorfields Eye Hospitals. Instruments were distributed online and via paper. Descriptive analyses were undertaken on completeness and time taken to complete. Initial exploratory analyses of the structural validity of the tool were undertaken, specifically, calculation of Cronbach’s alpha for each domain, (with an acceptable minimum value set at α = 0·70, and exploration of the impact of removing items on α)^29^, inter-item and item-total score correlation within domains (using Spearman’s rank correlation coefficients) and correlation between domains. Analyses were undertaken using Microsoft Excel and STATA (version 15.1, StataCorp).

## 3. Results

A total of 113 children and 57 parents/carers (table 1) participated in item development (figure 1). Participant demographics for those who took part in the qualitative phases of research (Phases 1 and 3) and in item assessment (Phase 4) are presented in table 1.

**Figure 1.**
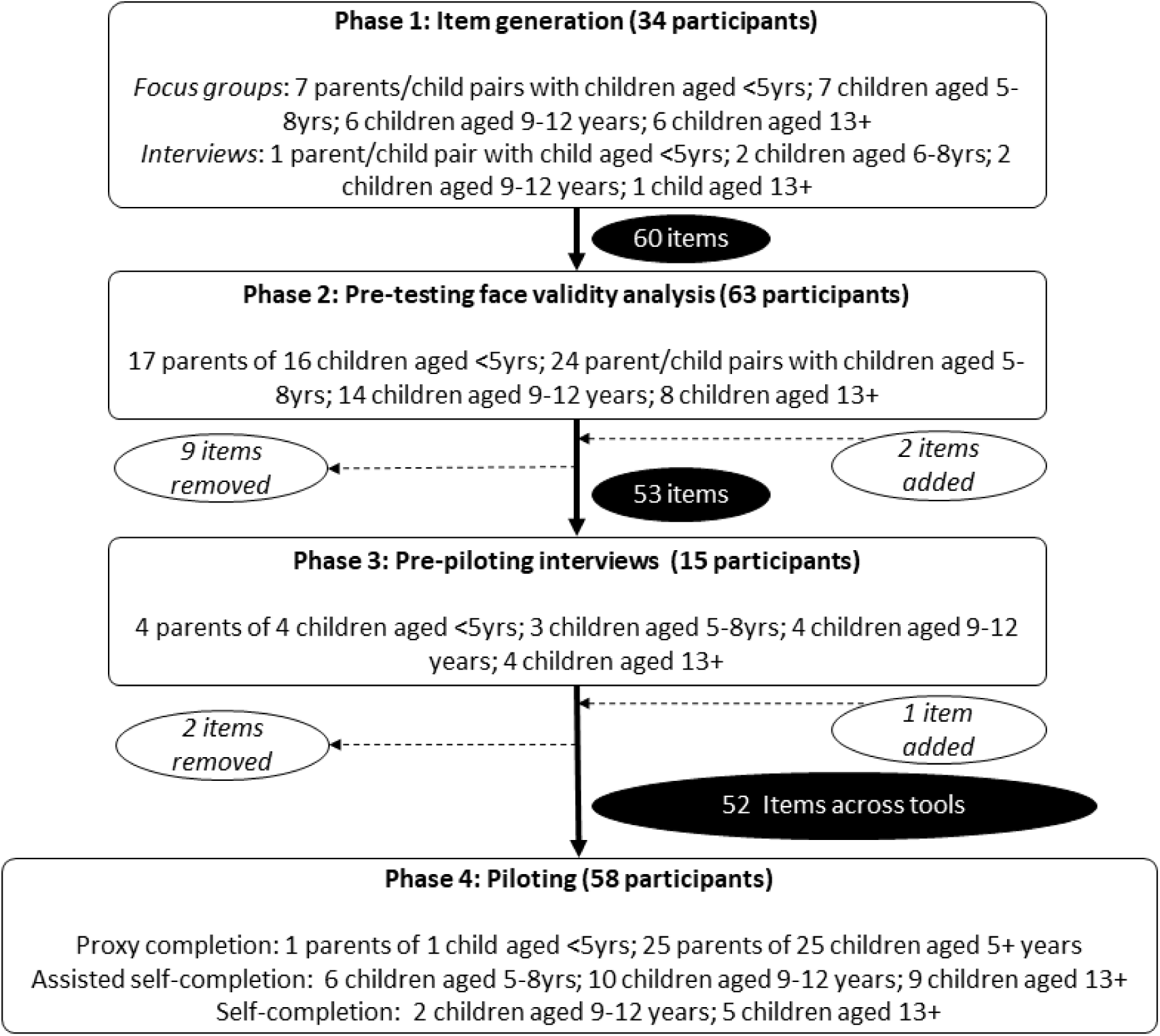
Participant flow chart.

**Table 1.** Family level participant characteristics.

| *Characteristic* | Phase 1 and Phase 3<br>(interviews / focus groups)<br>n=49 | Phase 4<br>(piloting)<br>n=58 |
| --- | --- | --- |
|  | n (%) | n (%) |
| *Ethnicity of respondent* |  |  |
| White - English, Scottish, Welsh, Irish | 16 (32·7) | 38 (65·5) |
| White other | 5 (10·2) | 5 (8·6) |
| Black African | 3 (6·1) | 2 (3·4) |
| Black Caribbean | 1 (2·0) | 1 (1·7) |
| Black Other | 2 (4·1) | 1 (1·7) |
| Indian | 2 (4·1) | 3 (5·2) |
| Pakistani | 2 (4·1) | 2 (3·4) |
| Bangladeshi | 2 (4·1) | 0 |
| Chinese | 1 (2·0) | 0 |
| Asian other | 1 (2·0) | 1 (1·7) |
| Other | 3 (6·1) | 1 (1·7) |
| Preferred not to say | 3 (6·1) | 4 (6·8) |
| Missing | 8 (16·3) | 0 |
| *Gender of child* |  |  |
| Female | 23 (46·9) | 26 (44·8) |
| Missing | 0 | 0 |
| *Family structure* |  |  |
| Two parent family | 25 (51·0) | - |
| Single parent family | 13 (26·5) | - |
| Other | 3 (6·1) | - |
| Missing | 8 (16·3) | - |
| *Highest education level of parent* |  |  |
| Non-university graduate | 10 (29·4) | - |
| Graduate | 26 (76·4) | - |
| Postgraduate | 5 (10·2) | - |
| Missing | 8 (16·3) | - |
| *Home ownership status of parents* |  |  |
| Owned | 22 (44·9) | - |
| Privately rented | 11 (22·4) | - |
| Rented from a housing association or local authority | 6 (12·2) | - |
| Other | 2 (4·1) | - |
| Missing | 8 (16·3) | - |

### 3.1. Phase 1

Phase 1 (figure 1) resulted in the generation of 60 items around characteristics (19 word items and 13 image items for the self-completion tools), severity (five items), exacerbators (five), location (six), temporality (three), and external presentation of symptoms (two), and proxy report (seven items). These items considered either qualitative or quantitative aspects of sensations and symptoms.

Other comments from participants ranged from accepting of instrument utility “*Because sometimes you can get a bit shy around people, and just get a bit silent, so having {the tool} to show is really good*”{child/C, 9-12 years}, and “*maybe that question would have made me go - Oh, yeah, actually, yeah, why is x doing that*”{parent/P}, to uncertainty about utility: “*I just think somebody like x who is pretty good at not really noticing pain, {so} it’s not terribly helpful*”{P} and “*I would…just talk to the doctor and my mum about how my eyes are feeling*” {C,9-12 years}.

### 3.2. Phase 2

Survey responses were received from 41 parents of affected children and 46 children and young people (with 22 completing the survey alone) (figure 1). Seven items did not reach the threshold for retention, and were flagged for potential removal from the item pool. The threshold for action was also reached for similarities between the terms “Hot” and “Burning”; “Itch” and “ Scratch”; and “Throbbing” and “Pulsating” within the characteristic domain. The majority of child and young persons who provided a preference on timing of administration chose the ‘any time needed’ option (13/19, 68%). Three additional items were generated in the temporality domain. Most of respondents (39/63) selected ‘Both online and paper’ availability of the future tool. Rephrasing was not suggested for any item. Rewriting and modification of retained items resulted in a tool for children aged 6 years and older, with self-completion and assisted completion options, and a proxy completion tool for younger children.

At the meeting of the Council of expertise, all terms within the tools were judged to be relevant to conditions other than uveitis by at least one member (i.e., no items were removed). The terms thought to be indicative of conditions other than uveitis by the majority were ‘Itch’ (characteristic domain) as selected by 67% (6/9) and ‘My child is avoiding looking at picture or words in books’ (proxy domain, 56%, 5/9). The Council agreed by consensus with the decision to merge the terms ‘Hot/Burning’, ‘Itch/Scratch’ and ‘Throbbing/Pulsating’ rather than select only one for use.

The resultant self-completion and assisted completion tools comprised: three questions in the characteristic domain, with answer options covering the 14 word based and 9 image based items; two questions (two items) in the severity domain, one question (six items) in the temporality domain, one question (four items) on exacerbators, three questions (eleven items) on location, and two questions (two items) in the external presentation domain. Assisted completion was defined and communicated to participants as an adult parent or carer helping the child to access the instrument, and or to read the questions, but allowing the child freedom of response, with guidance that questions should be left unanswered if the child was unable to understand it without an explanation from the adult. The proxy completion score had seven items. Both tools also had summary overall questions (‘My eyes feel exactly how I want them to feel for self-completion’, and ‘I am sure that my child’s eyes are comfortable for them’) answered using a visual analogue scale (running from ‘Completely comfortable’ to ‘Worst they have ever felt’ / ‘Extremely uncomfortable for them’. Where other quantitative responses were needed, a four level Likert scale ^30^ was used: ‘not at all true’, ‘a little bit true’, ‘mostly true’ and ‘completely true’.

### 3.3. Phase 3

Following the pre-piloting interview (figure 1), none of the 15 participants suggested changes in terms of mode of administration. All the eight children aged over 8 years (range 9 – 15 years old) were able to self-complete, and one of the three children aged 5-8 years self-completed the tool without assistance. One question on external presentation (inviting respondents to annotate images of the upper face to show where there was any eye redness) had a low rate of completion (not completed by any respondent) and was removed with retention of a question allowing annotation of a closer-up image of the eye. One additional item, assessing acceptability and user experience, was suggested either directly or indirectly by three respondents (e.g., “*It feels funny to talk about my eyes like this*”) {C,9-12 years}).

Following re-modification of the retained items, the two resulting self-completion and assisted self-completion tools for children aged 9 years and older, and those aged 5 years and over respectively, each comprised 16 questions, covering 46 items: One group of questions combining the characteristic and severity domain, with answer options covering the 14 word based and 9 image based items; one question (six items) in the temporality domain; four questions (four items) on exacerbators; three questions (nine items) on location, and one question (one item) in the external presentation domain. An additional question was added to the self-completion tool to gauge acceptability: “How did telling us about your eyes make you feel?” with a visual analogue response scale running from 0 (‘It made me feel worse’) to 10 (‘It made me feel better’) with a midpoint of 5 (‘It doesn’t make me feel worse or better’).

The proxy completion score, for parents and carers, had seven items. Both tools also had summary overall scores for eye sensations and symptoms completed using visual analogue scoring (running from 0 to 10).

### 3.4. Phase 4

A total of 60 participants started the tool, of whom only two failed to complete (in both cases the child was symptom free). Of the 58 children or parent/carers who completed the tool, there was proxy completion by 26 parents (for 1 child aged under 5 years and 25 children aged 6+ years), assisted self-completion by 25 children aged 5+ years, and self-completion by 7 children aged 10 to 15 years (figure 1). Time taken to complete ranged from 2 to 28 mins (median time 8 mins) for self-completion and 1 to 11mins 20secs (20secmedian 5 mins) for proxy completion.

Descriptive analyses of responses showed high levels of completion for all items (table 2 and 3). Four respondents (4/26, 3.9%) chose to submit an annotated image of the affected eye. Inter-item correlation within domains (supplemental tables 1 and 2) suggested overly high (>0·5 threshold)^29^ correlations between the items measuring exacerbation with sunlight, eye movement and reading in the self-completion tool, and between blinking and keeping eyes closed in the proxy measurement tool.

**Table 2.**
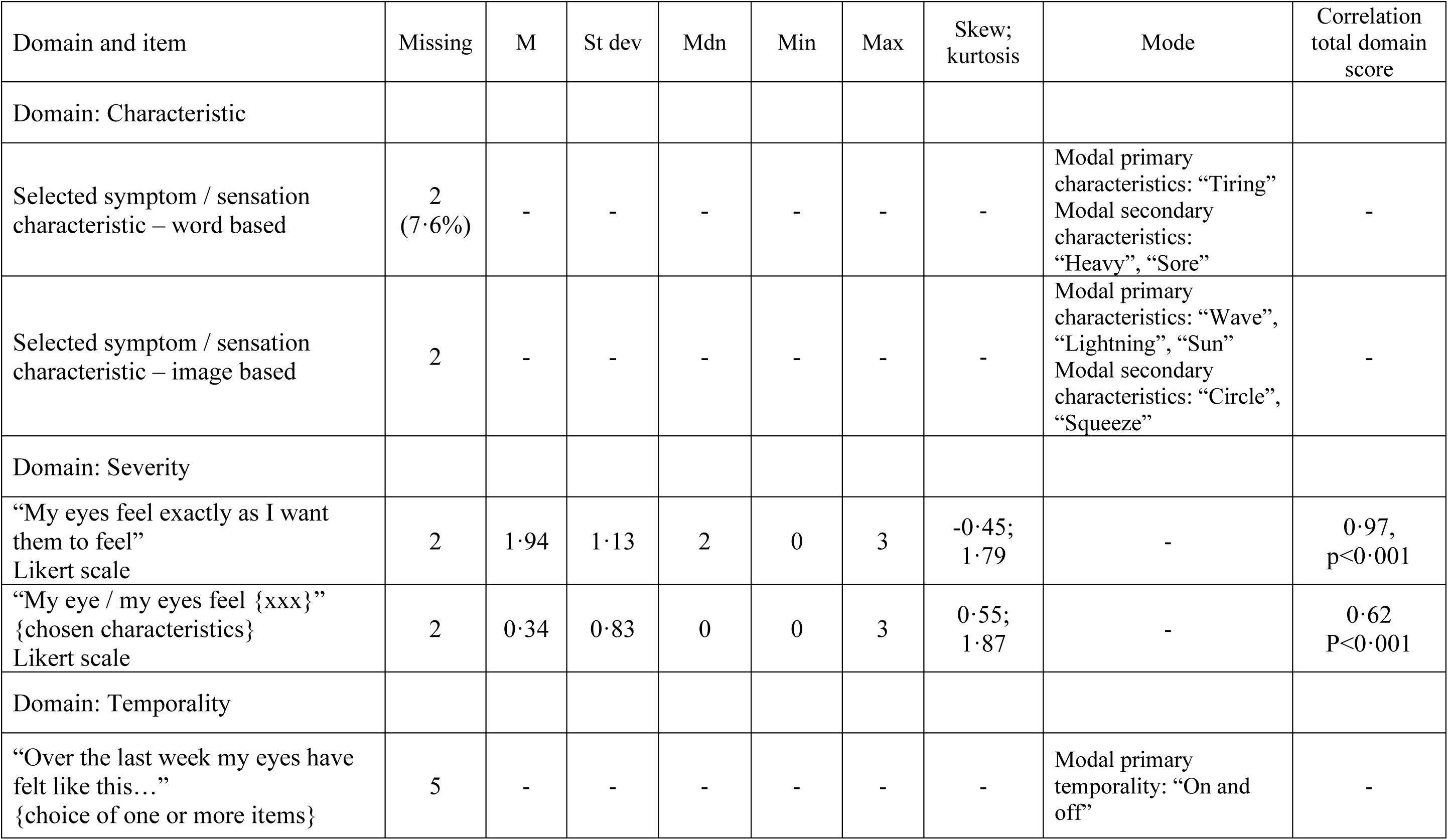
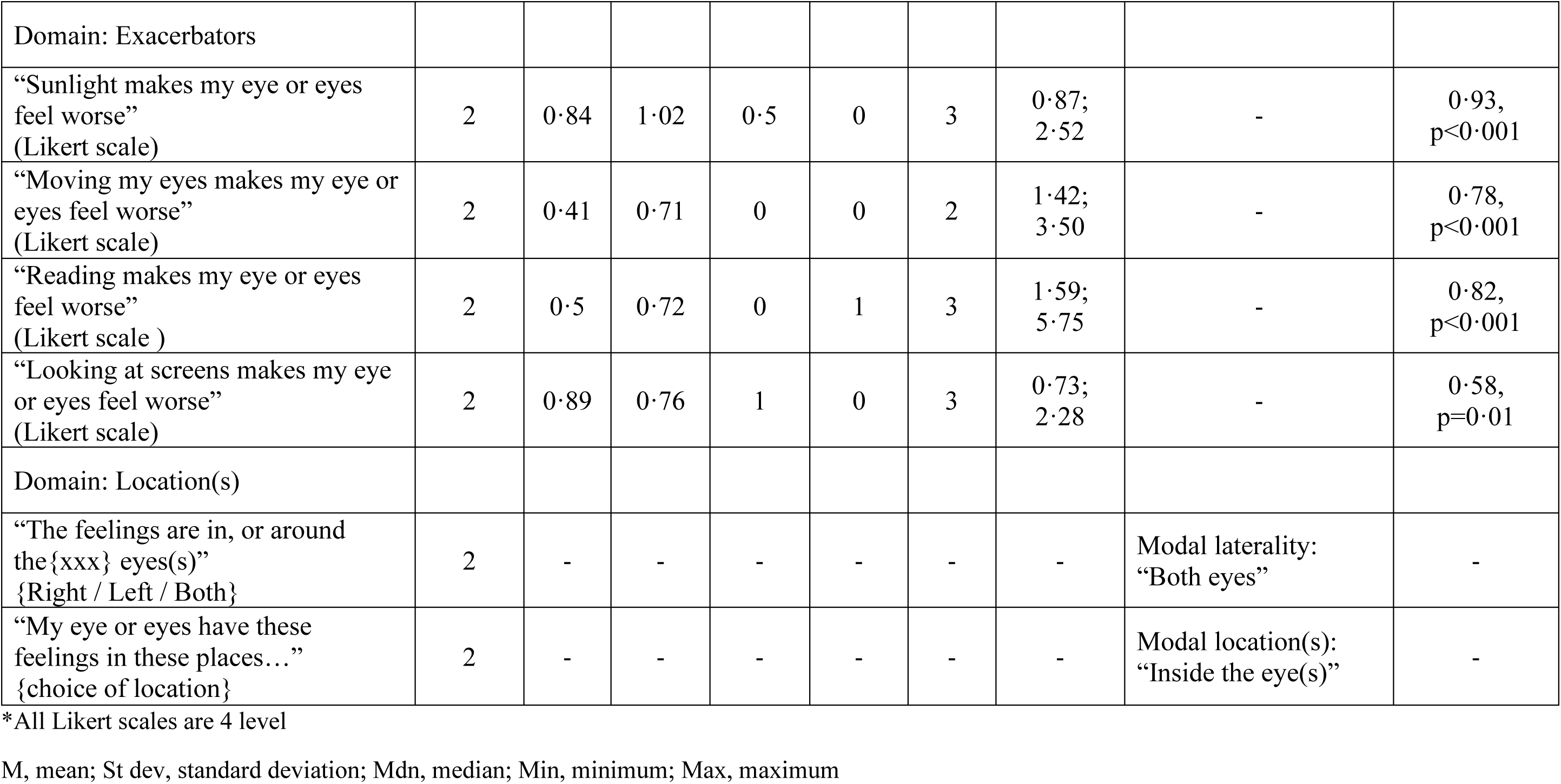
Response rates and scores for self-completion tool items*.

| Domain and item | Missing | M | St dev | Mdn | Min | Max | Skew;<br>kurtosis | Mode | Correlation<br>total domain<br>score |
| --- | --- | --- | --- | --- | --- | --- | --- | --- | --- |
| Domain: Characteristic |  |  |  |  |  |  |  |  |  |
| Selected symptom / sensation<br>characteristic – word based | 2<br>(7.6%) | - | - | - | - | - | - | Modal primary<br>characteristics: “Tiring”<br>Modal secondary<br>characteristics:<br>“Heavy”, “Sore” | - |
| Selected symptom / sensation<br>characteristic – image based | 2 | - | - | - | - | - | - | Modal primary<br>characteristics: “Wave”,<br>“Lightning”, “Sun”<br>Modal secondary<br>characteristics: “Circle”,<br>“Squeeze” | - |
| Domain: Severity |  |  |  |  |  |  |  |  |  |
| “My eyes feel exactly as I want<br>them to feel”<br>Likert scale | 2 | 1.94 | 1.13 | 2 | 0 | 3 | -0.45;<br>1.79 | - | 0.97,<br>p<0.001 |
| “My eye / my eyes feel {xxx}”<br>{chosen characteristics}<br>Likert scale | 2 | 0.34 | 0.83 | 0 | 0 | 3 | 0.55;<br>1.87 | - | 0.62<br>P<0.001 |
| Domain: Temporality |  |  |  |  |  |  |  |  |  |
| “Over the last week my eyes have<br>felt like this...”<br>{choice of one or more items} | 5 | - | - | - | - | - | - | Modal primary<br>temporality: “On and<br>off” | - |
| UveQ development |  |  |  |  |  |  |  |  |  |
| Domain: Exacerbators |  |  |  |  |  |  |  |  |  |
| “Sunlight makes my eye or eyes feel worse”<br>(Likert scale) | 2 | 0.84 | 1.02 | 0.5 | 0 | 3 | 0.87;<br>2.52 | - | 0.93,<br>p<0.001 |
| “Moving my eyes makes my eye or eyes feel worse”<br>(Likert scale) | 2 | 0.41 | 0.71 | 0 | 0 | 2 | 1.42;<br>3.50 | - | 0.78,<br>p<0.001 |
| “Reading makes my eye or eyes feel worse”<br>(Likert scale ) | 2 | 0.5 | 0.72 | 0 | 1 | 3 | 1.59;<br>5.75 | - | 0.82,<br>p<0.001 |
| “Looking at screens makes my eye or eyes feel worse”<br>(Likert scale) | 2 | 0.89 | 0.76 | 1 | 0 | 3 | 0.73;<br>2.28 | - | 0.58,<br>p=0.01 |
| Domain: Location(s) |  |  |  |  |  |  |  |  |  |
| “The feelings are in, or around the{xxx} eyes(s)”<br>{Right / Left / Both} | 2 | - | - | - | - | - | - | Modal laterality:<br>“Both eyes” | - |
| “My eye or eyes have these feelings in these places...”<br>{choice of location} | 2 | - | - | - | - | - | - | Modal location(s):<br>“Inside the eye(s)” | - |

\*All Likert scales are 4 level
M, mean; St dev, standard deviation; Mdn, median; Min, minimum; Max, maximum

**Table 3.** Response rates and scores for proxy-completion tool items*.

| Item | Missing | M | St dev | Mdn | Min | Max | Skew; kurtosis | Correlation total domain score |
| --- | --- | --- | --- | --- | --- | --- | --- | --- |
| “I am sure that my child’s eyes are comfortable for them”<br>(Likert scale) | 0 | 1.89 | 0.91 | 2 | 0 | 3 | -0.42;<br>2.42 | 0.69,<br>p<0.001 |
| “My child is more irritable”<br>(Likert scale) | 0 | 0.31 | 0.55 | 0 | 1 | 2 | 1.55;<br>4.46 | 0.39, p=0.05 |
| “My child is rubbing their eyes more often”<br>(Likert scale) | 0 | 0.61 | 0.80 | 0 | 0 | 2 | 0.79;<br>2.06 | 0.84,<br>p<0.001 |
| “My child is blinking their eyes more often”<br>(Likert scale) | 0 | 0.19 | 0.40 | 0 | 0 | 1 | 1.56;<br>3.43 | 0.46, p=0.02 |
| “My child is avoiding light”<br>(Likert scale) | 0 | 0.77 | 1.11 | 0 | 0 | 3 | 1.19;<br>2.96 | 0.84,<br>p<0.001 |
| “My child is avoiding looking at pictures or words in book”<br>(Likert scale) | 0 | 0.11 | 0.33 | 0 | 0 | 1 | 2.41;<br>6.80 | 0.43, p=0.03 |
| “My child is wanting to keep their eyes closed”<br>(Likert scale) | 0 | 0.08 | 0.27 | 0 | 0 | 1 | 3.18;<br>11.08 | 0.48, p=0.02 |
| “My child is avoiding screens”<br>(Likert scale) | 0 | 0.08 | 0.27 | 0 | 0 | 1 | 3.18;<br>11.08 | 0.40, p=0.04 |

\*All Likert scales are 4 level

Correlations scores between individual item and whole domain scoring were consistently above 0·58 for the self-completion tools (meeting the threshold of 0·3 for correlation coefficients often set for psychometric evaluation)^29^ and above 0·39 for the proxy completion tool (table 4). Initial estimation of Cronbach’s alpha for the tool overall was 0·84, with within-domain alphas of 0·81 – 0·87. The proxy completion tool scored α=0·69, with removal of the items measuring ‘blinking’ and ‘irritated’ improving structural validity (α=0·74, supplemental tables).

**Table 4.** Internal consistency scores for self and proxy completion tools.

| Tool | Domain | Items and scoring schema | Phase 4 scoring results | Domain Cronbach alpha* |
| --- | --- | --- | --- | --- |
| Self- / Assisted self-completion |  |  |  |  |
|  | Severity | 2 items, score range: 0 – 3<br>Total score range 0 – 6 | Median total score 1, IQR 0 – 3, range 0 – 6<br>Mean 1.4, std dev 1.7, skewness 1.3, kurtosis 4.0 | 0.87 |
|  | Exacerbators | 4 items, score range 0 – 3<br>Total score range 0 – 12 | Median total score 1, IQR 0 – 4, range 0 – 8<br>Mean 1.8, std dev 2.1, skewness 1.0, kurtosis 3.5 | 0.81 |
| Proxy completion |  |  |  |  |
|  | Proxy items | 7 items, score range 0 – 3<br>Total score range 0 – 21 | Median total score 1, IQR 1 – 4, range 0 – 8<br>Mean 2.2, std dev 2.4, skewness 0.9, kurtosis 2.8 | 0.69<sup>‡</sup> |

\*As a measure of internal consistency
<sup>‡</sup>impact of item removal: removal item 1 $\alpha=0.74$ ; item 2 $\alpha=0.63$ ; item 3 $\alpha=0.72$ ; item 4 $\alpha=0.61$ ; item 5 $\alpha=0.71$ ; item 6 $\alpha=0.68$ ; item 7 $\alpha=0.68$

## 4. Discussion

From this prospective, mixed-methods, multi-phase study we report the first stage development of a suite of age-appropriate novel patient reported outcome tools (for self and for proxy completion) using item generation and validation (face validity) for childhood ocular symptoms. These demonstrate the feasibility of capturing these sensations, with high acceptability and high rates of instrument completion. Early analyses suggest acceptable internal consistency for the tools.

This patient-centred development, with elements such as the use of visual based methods to enhance rapport and communication with child participants,^31^ has resulted in patient-reported disease activity metrics which may be useful as an outcome measure in disease monitoring and as a method of classifying disease to improve understanding of phenotype. Despite the negative experience, pain and other symptomatology can be of benefit to the individual, by triggering necessary health care seeking behaviour. Symptoms may also provide useful diagnostic information on disease entities, and help to deepen understanding of disease mechanisms. One such example is the absence of pain in ‘silent’ myocardial infarction, now recognised as a biomarker for autonomic dysfunction.^32,33^ Although the pathophysiology of pain is multimodal and complex, inflammatory signalling molecules are thought to play a central role in inflammatory pain mechanism. The absence of pain in some forms of childhood ocular inflammatory disease may be due to differences in disease mechanism – for example, children with spondyloarthropathies and/or those who carry *HLA-B*27,* may present with a red and/or painful eye, in a similar pattern to those with adult onset disease.^34^

For children with uveitis symptoms, either captured by the child themselves, or captured by carers observing signs of discomfort, may be an informative discriminator of cytokine profile, and thus disease mechanism and disease subtype beyond informing on *HLA-B*27* status. Another example from the broader medical literature is the different somatosensory phenotypes in peripheral neuropathic pain disorders of childhood and adolescence, with these phenotypes potentially providing a means to individualise therapeutic approaches.^35^

Paediatric pain and symptom experiences are under-represented within the pain literature, with recent calls to make pain in children “matter”, make it “understood”, make it “visible”, as well as making it “better”.^36^ Pain is particularly invisible in the context of childhood chronic disorders, and ‘visceral’ disorders.^36,37^ Childhood ocular inflammatory disease symptoms are therefore in particular need of attention. Existing eye pain scores have been designed to measure post-operative discomfort, or pain related to adult eye surface disorders^38^ rather than sensations linked to disorders occurring within the ‘viscera’ of the eye. Visceral sensations can be challenging to communicate. The use of pictograms to support children and young people in characterising and communicating their sensory experiences, although not novel within the wider field of pain metrics,^39,40^ are novel approaches within eye pain metrics. Our child driven approach to image development and use of a diverse pool of participants should, and has, supported the utility and representativeness of the selected images. The development of a behavioural symptom metric, anchored in explorations of family experience, has resulted in disease and organ specific items missing from other similar behavioural pain scales,^41^ and which again should support utility in this clinical area.

There are limitations around representativeness, and accessibility. The UK distribution of childhood uveitis by ethnicity and socioeconomic background is unclear. However, the sampling and recruitment approach has created a study population with representation of a wide range of patient backgrounds. Imaging based responses, although innovative and welcomed by patients, are inaccessible to children with impaired vision. This limitation does not prevent the use of this score for those children with visual disability, with the use of word based responses for capturing symptoms, and a quantitative scoring schema independent of the imaging based items. The instrument is also limited by the pragmatic decision to not involve children with additional communication barriers, e.g. neurodevelopment impairments or language barriers. The parent/proxy versions does however ensure that something of their experience might be captured. There is also evidence, from the Phase 2 Council of Expertise, of some clinical utility of the ChOIR-Q in ocular disorders beyond uveitis.

The use of Cronbach alpha to describe internal consistency is limited by the absence of an analysis of structural validity for this initial stage of development and validation of the tool, and limited by the relatively small number of children within subgroups as categorised by age and ethnic background. Future development of the instrument, following psychometric evaluation and demonstration of structural and clinical utility, will seek to consider how to involve diverse patient populations, as well as understanding performance in international patient groups, with a final aim to ensure a valid, robust instrument for use in clinical practice and medical research.

## Appendix A. Supplemental content

Supplement 1: Interview Topic Guide

## Appendix B. Supplemental content

Supplement 2: Supplemental table 1. Inter-item correlation for the exacerbator domain in the self-completion tool, and Supplemental table 2. Inter-item correlation for the proxy completion tool

## Author contributions

A.L.S. led the study conceptualization and design; A.L.S and V.T. co-led the development of methodology, data curation and analysis. A.L.S. led the formal analysis, visualization, and initial manuscript drafting and editing. J.S.R. supported design. All authors supported interpretation of results, all authors assisted with manuscript editing, all authors approved the final manuscript as submitted.

## Data Availability

Individual level data are not being made available for this study involving human research participant data. Consent for publication of raw data was not obtained (as this approach may have led to differential non-inclusion of certain patient groups). The dataset could in theory pose a threat to confidentiality. Guidance was sought from relevant Ethics Committee.

## Acknowledgments

We are grateful to all of the study participants and their families: this work was made possible by their contributions. We thank Lis Cordingley, Senior Lecturer in Health Psychology, Division of Musculoskeletal and Dermatological Sciences, University of Manchester, and Dr Elena Moraitis, Consultant Rheumatologist, Great Ormond Street Hospital, for their input, in particular their contribution to the Council of Expertise. We thank the Childhood Uveitis Study Steering patient group for their contribution to study design and implementation, and for their contribution to the Council of Expertise. We thank the United Kingdom Paediatric Ocular Inflammatory Group for their support with patient recruitment.

